# Enhancing Interventional Cardiology Training: A Porcine Heart-Based Coronary Intervention Simulator

**DOI:** 10.1101/2024.09.02.24312944

**Authors:** Joerg Reifart, Paul Anthony Iaizzo

**Affiliations:** Visible Heart Laboratories, Department of Surgery, University of Minnesota, Minneapolis, Minnesota, USA

## Abstract

**Introduction:** Access to simulators for interventional cardiology is currently limited. High acuity, low occurrence procedures (HALO), such as coronary perforation or iatrogenic dissection, are not trained in currently available simulators. We developed a cost-effective coronary intervention simulator designed to enhance the training of both novice and experienced interventionalists.

**Methods:** Porcine hearts from 6-month-old Yorkshire pigs (heart weight: 300-500g) were obtained from a large animal research laboratory. Guide catheters were inserted into the coronary artery ostia and secured with superglue. To maintain shape and rigidity, commercially available polyurethane insulation foam was injected into the ventricles. The guide catheter was then connected to a Tuohy valve linked to a 3-way stopcock. One connection led to a pressure infusion bag filled with tap water (inflated at > 300 mmHg); the other was used for contrast injection. The heart was set on a radiolucent box with a fluid collector underneath. Clinical scenarios were simulated using 3D-printed stenoses and occlusions, blood clots for occlusive myocardial infarction, balloon oversizing for dissections, and needle trauma for perforations.

**Results:** The simulator was used to practice coronary angiography, managing perforations, bifurcations, dissections, and acute coronary occlusions.

Assembly, set-up, and simulation time until refilling the perfusion bag was required were 50, 25, and 30 minutes, respectively. Intravascular imaging with Optical Coherence Tomography was successfully used to guide interventions. The simulator was frozen and reused more than three times without notable deterioration.

**Conclusion:** A wide range of clinical scenarios can be trained with our model. Its preparatory flexibility, including the ability to be frozen for on-demand training, enhances its utility. Limitations include the absence of pulsatile flow, heart movement, and the inability to train guide catheter intubation.

## INTRODUCTION

Several studies showed the benefits of simulation of interventional or angiographic training with virtual reality simulators to have new interventional fellows gain their first experience with coronary procedures[1–3]. Hands-on alternatives are physical computer-based simulator systems, which have learners more practically experience angiography and coronary interventions[4–7]. However, because these systems are cost-prohibitive (only available online quote: $55.380), this type of training is not currently widely adopted[8].

Most interventional trainees take their first interventional steps, learning angiography and simple procedures in patients under close supervision by their respective mentors. This may increase patient risk, but it is also associated with high psychophysiologic stress experienced by the trainees[9].

Here, we present a low-cost system using porcine hearts that could be conjured from large animal research facilities or possibly a slaughterhouse to create a training system. It allows novice interventionalists to learn both what angulations to use in coronary angiography as well as coronary interventions, including complication management training.

## METHODS

### HEART PREPARATION

Porcine hearts (fresh or frozen) from 6-month-old Yorkshire pigs (weight animal 60 – 80kg, weight of the hearts 300 -500g) that had been used for differing research projects in the Visible Heart Laboratories were used in this model.

Depending on the extent of dissection, guide catheters (6-8F JL4.0, EBU 3.75, or JR 4.0—previously used) were placed into both coronary ostia (Figure 1, (C)). The aortic root can be trimmed in case of excess aortic tissue or difficulty canulating the ostia. To prevent the catheters from exiting the ostia, they were fixed in position with super glue.

**Figure 1:**
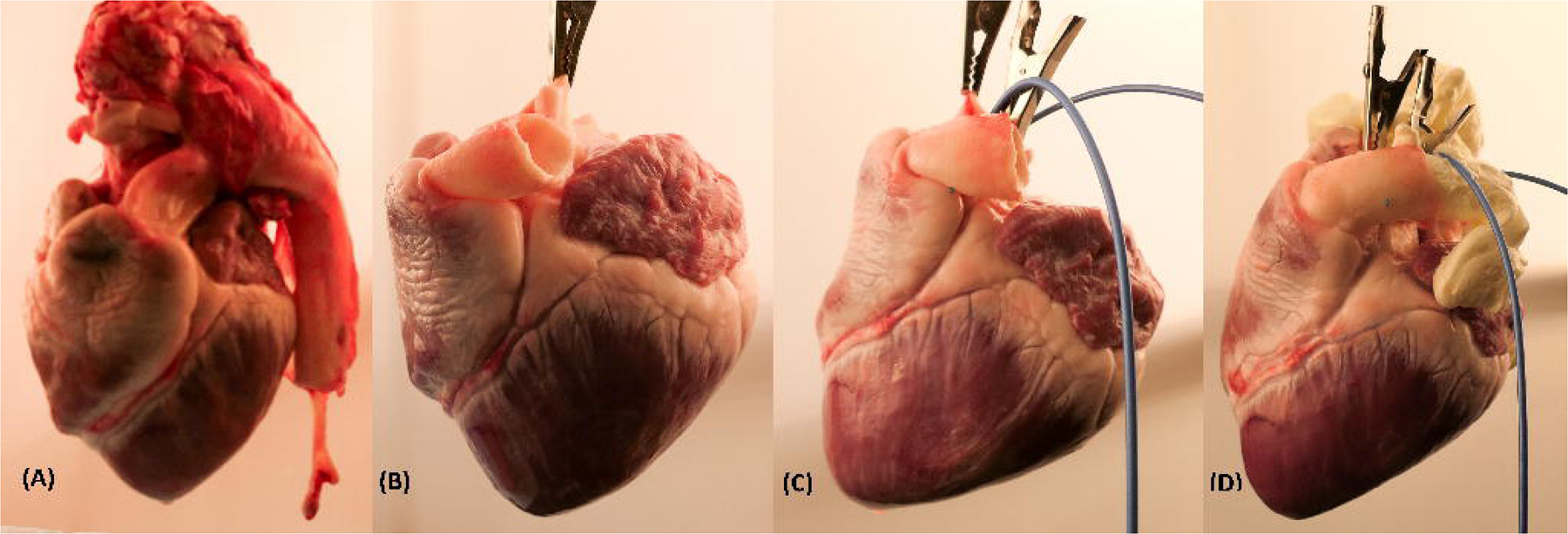
(A) Porcine heart with aorta attached. (B) Heart after cutting the Aorta so the coronary ostia can be accessed easily. (C) Guide catheters placed into the right and left coronary ostium and fixed in place with Super-glue. (D) Injection of polyurethane foam into the left and right ventricle, leading to expansion of all cavities and great vessels.

We used commercially available polyurethane insulation foam (6.99$) to fill the ventricles, atria, the pulmonary artery, and the aorta. This is to prevent fluid from entering the ventricles, fix the guide catheters in place, and to lend support to the organ, resulting in realistic coronary projections (Figure 1 (D)). Excess insulation foam was cut. Foam in hard-to-reach places or within vessels can be dissolved using commercially available acetone. The guide catheter was then connected to a Tuohy valve and a manifold, of which one port was connected to a pressure infusion bag filled with tap water (Figure 2; Supplemental Digital Content 1 - Figure for fast refilling method). System assembly duration was timed in minutes. The total cost for the used materials was $42.56 (Table 1).

**Figure 2:**
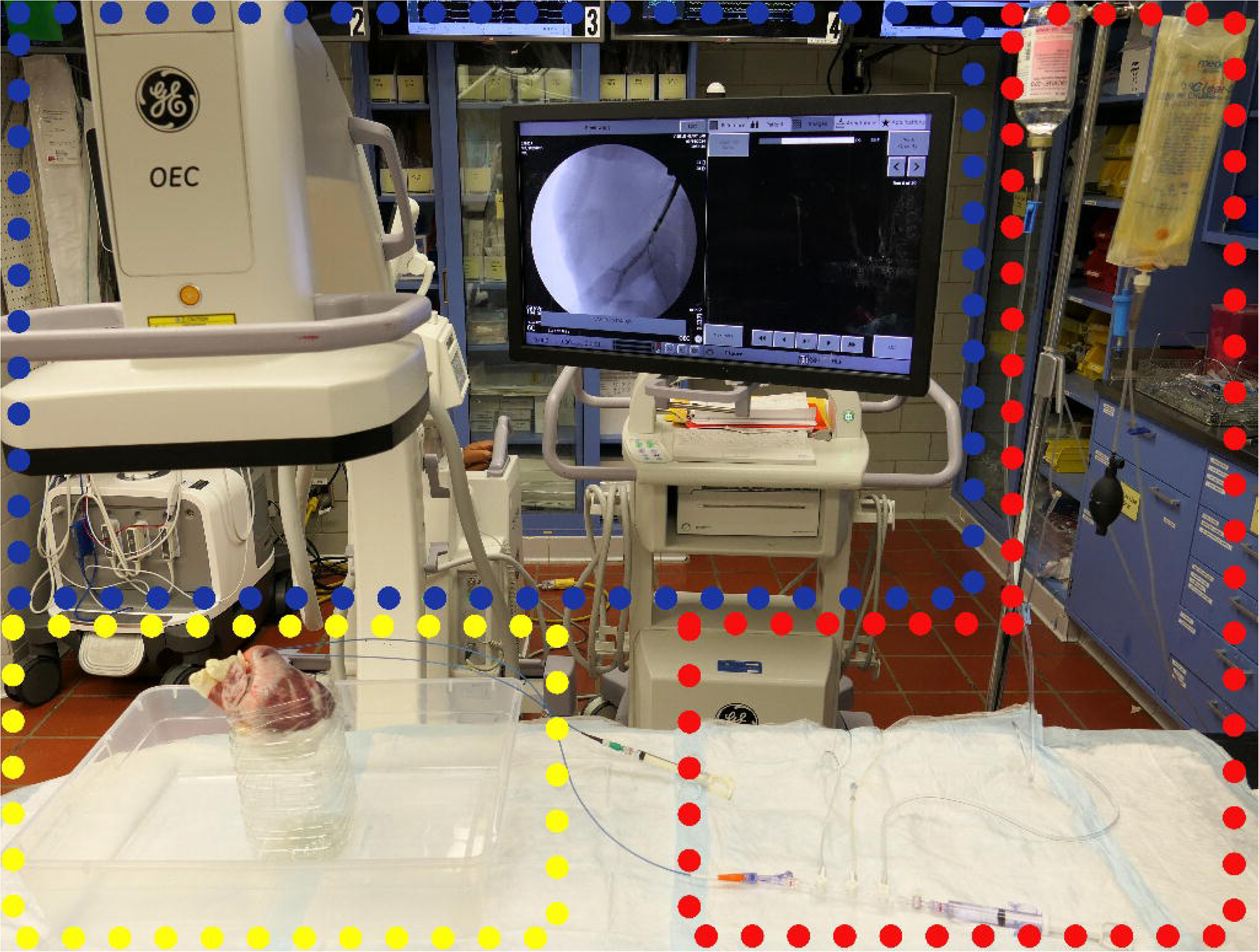
Coronary Intervention Simulator final setup. Fluoroscopy C-Arm on the left (blue box). Prepped heart with guide catheters in place below the detector on the support structure putting it at the isocenter of the fluoroscopy machine and allowing to follow all clinically used coronary projections (yellow box). Pressure infusion set on the right, as well as contrast solution – following a standard catheterization laboratory setup (red box).

**Table 1:**
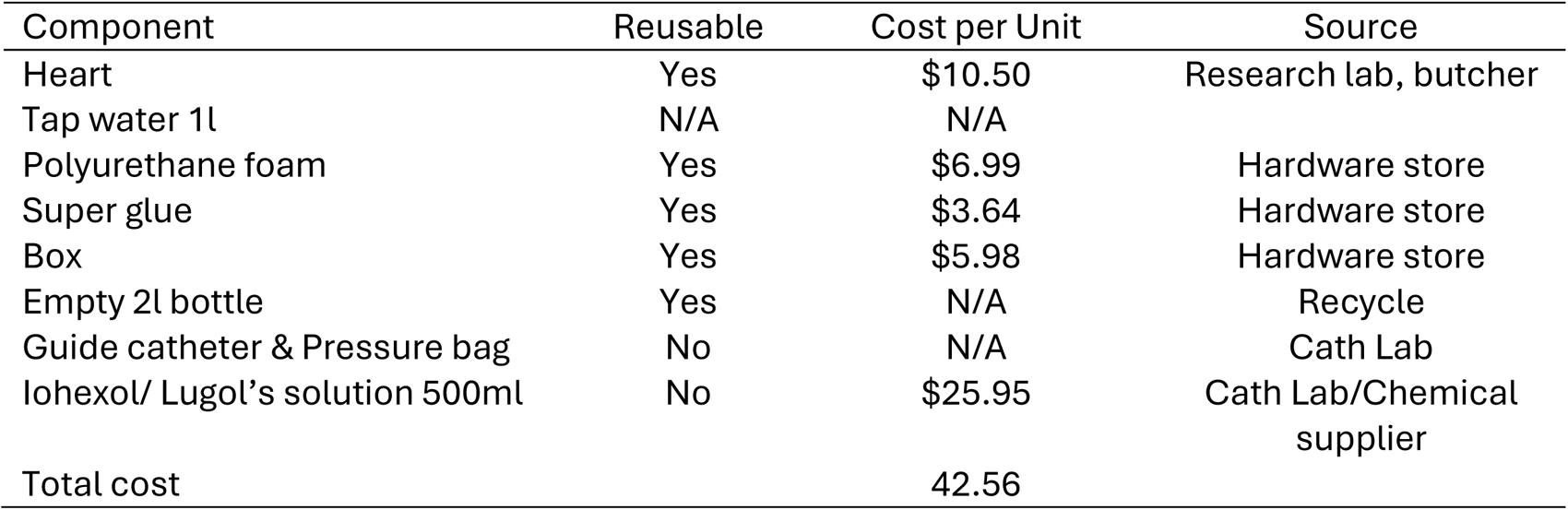
Cost of Materials Used for single reusable PCI Trainer.

### SYSTEM SET-UP

The heart was placed on a support structure for which either a clear 28 Qt|27l Storage Box with a cut-off 2 liter PET bottle was fixed to the bottom with epoxy (Figure 2) or alternatively two Polyethylene terephthalate (PETE) packages from Affinity Fusion Oxygenators (Supplemental Digital Content 2 -suppl Figure). The support structure collects the fluid and contrast that circulates through the heart and positions it at a height and angle similar to in-human coronary angiograms.

The pressure infusion system was inflated to pressures ∼300mmHg, which allows for continuous antegrade contrast clearance when valves are opened (standard). If feasible in the local catheterization laboratory environment, a hose-based continuous perfusion set-up can be chosen to allow for uninterrupted training (Supplemental Digital Content 3 -Suppl Figure 3). We acquired residual unused Iohexol from Catheterisation Laboratories and Radiology departments for intravenous contrast.

Alternatively, 10% Lugol’s solution (Iodine potassium iodide) can be used as an inexpensive alternative contrast medium in this model. Lugol’s solution, however, will stain the materials used.

System set-up duration was timed in minutes.

### TRAINING SCENARIOS

Different standard training scenarios apart from standard angiography angulations, balloon/stent exchange practice, and handling of devices such as Optical Coherence Tomography (OCT) catheters for intravascular imaging were simulated:

- Circular sutures around the artery produced Simple Type A lesions.
- Thrombotic acute occlusions were simulated by injecting previously collected coagulated porcine blood directly before guide catheter placement or through the guide catheters themselves.
- Complex lesions like chronic totally occluded coronary artery lesions or severely calcified lesions were modeled from diseased human coronary arteries and then 3-D printed. In short: Formalin-fixed human hearts underwent iodine-enhanced micro-computed tomography scanning (North Star Imaging X3000 Micro-CT scanner, Rogers, Minnesota)) and were subsequently segmented (Figure 3, MIMICS 26.0, Materialise, Plymouth, MI, USA), Figure 3, A). The segmented coronary lesions were then 3-D printed at actual scale (Figure 3, B) and inserted into the coronary arteries before fixing the guide catheters to the coronary ostia.
- Short, proximal, calcified stenoses were created by injection of Super-Glue mixed with Barium sulfate powder into the artery before inserting the guide catheter.
- Dissections were created by inflating grossly oversized balloons (e.g., 4.5mm Balloon in a 2.5mm diameter vessel).
- Simple lacerations with a needle or scalpel were used to simulate coronary perforations.

**Figure 3:**
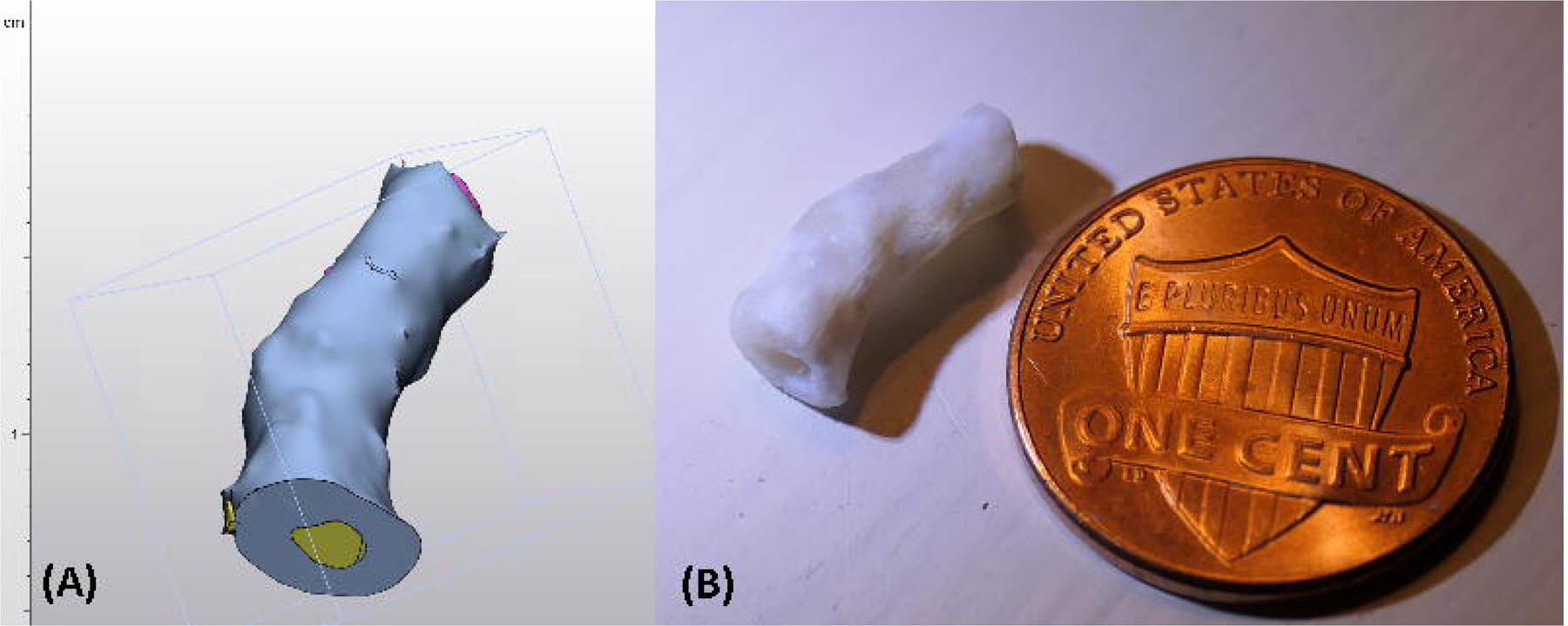
(A) Digital 3-D model of a coronary chronically occluded artery segmented from a high-resolution micro-computed tomography scan. (B) 3D print of the artery segment before introduction into the intervention simulator. Penny for scale.

Duration, until the perfusing pressurized saline bag had to be refilled, was timed in minutes. Further recommendations are in the supplementary Appendix.

## RESULTS

Initial assembly, including the polyurethane curing time of the coronary intervention trainer, was 50 minutes. Set-up in the catheterization laboratory was 25 minutes. Continuous antegrade perfusion on 1l pressure perfusion allowed for 30 minutes of uninterrupted simulation. High-pressure infusion through the guiding catheter via the connected pressured fluid bag resulted in appropriate, non-collapsed vessel diameters during interventions, with antegrade contrast clearance (see Videos, Supplemental Digital Content 4 & 5). Optical coherence tomography (OCT) worked to acquire intravascular images without the need for additional contrast injection.

Previously described scenarios were successfully simulated:

- Figure 4 : Angiographic and OCT image of the created acute thrombotic occlusion within the simulator.
- Figure 5 & Video, Supplemental Digital Content 6: Coronary perforation on OCT and angiography
- Video, Supplemental Digital Content 7: Angiography of the 3D-printed CTO.

**Figure 4:**
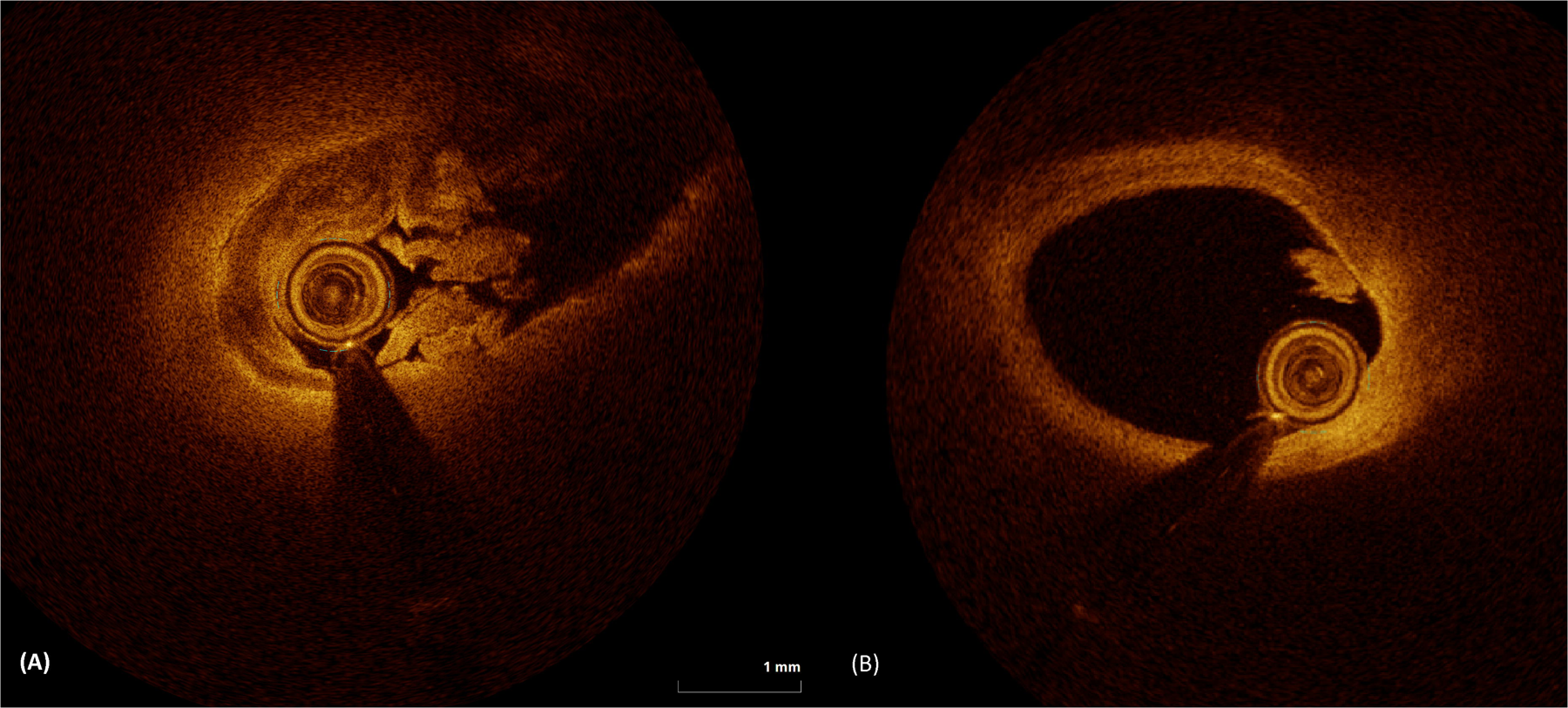
OCT showing occlusive (A) and non-occlusive (B) white thrombus in the RCA that was saved from a previous porcine case and then injected via the guide catheter. To train thrombus aspiration.

**Figure 5:**
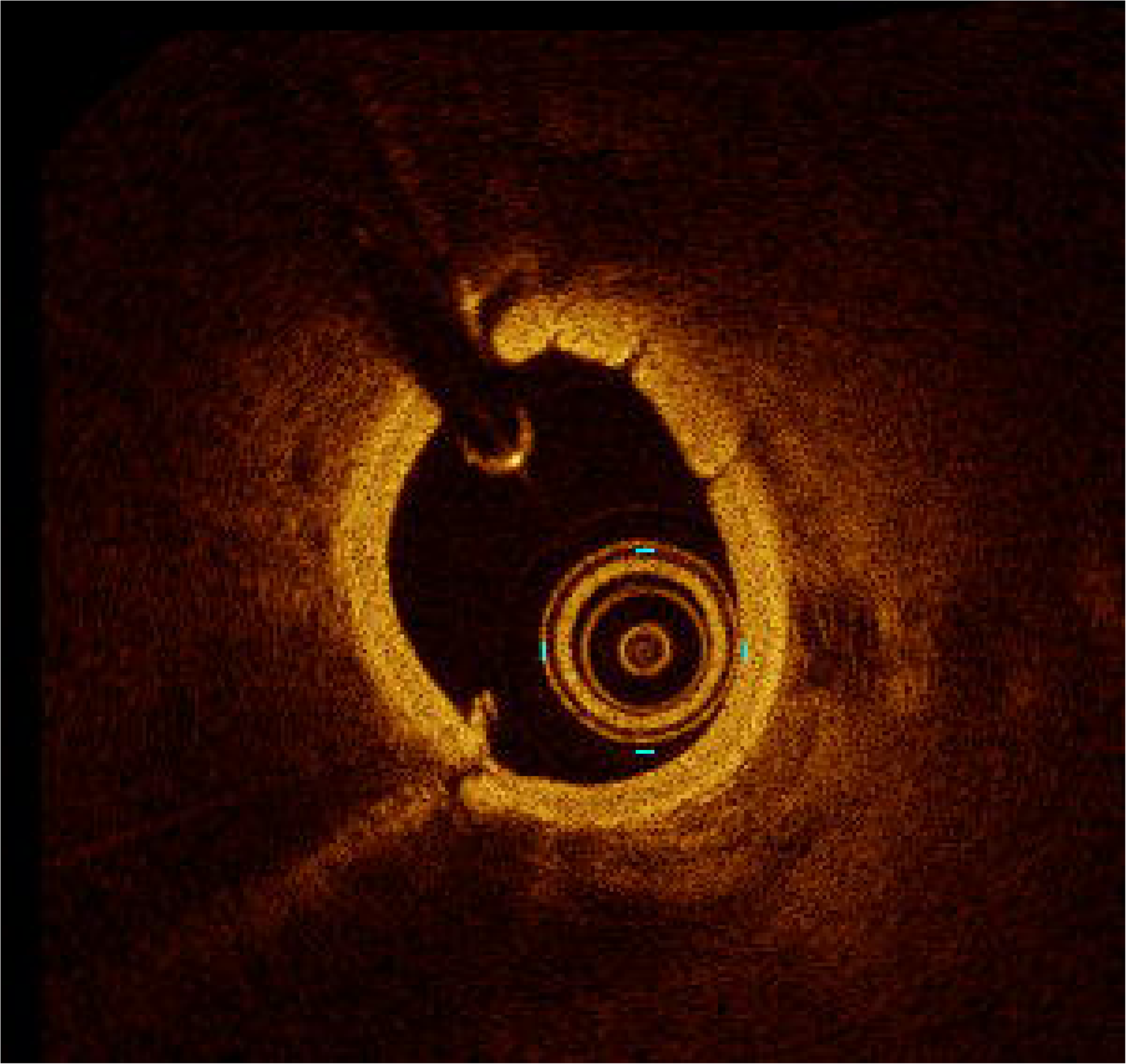
OCT of Ellis III coronary artery perforation before treatment.

Other trainable coronary interventions and maneuvers are presented in Table 2.

**Table 2:** Trainable and non-trainable clinical situations.

| **Trainable** | **Non-trainable** |
| --- | --- |
| Wiring of tortuous vessels and side-branches | Ostial Interventions |
| Wiring septal/other collaterals for retrograde approach | Bypass Graft interventions |
| Precise placement of stents in bifurcated lesions | Fractional Flow Reserve (FFR) |
| Transcoronary ablation of septal hypertrophy (TASH) | Electrophysiologic Interventions |
| Embolization to occlude perforated, small vessels | Structural Interventions |
| Placement of covered stents to seal perforations |  |
| Use of Atherectomy devices (rotational and orbital) |  |
| Use of OCT and IVUS to guide PCI* |  |
| Thrombus aspiration |  |
| *Optical Coherence Tomography (OCT) and Intravascular Ultrasound (IVUS) |  |

The assembled hearts with guide catheters in place were frozen and used again more than three times, with the main limitation to re-freezing and re-using being the accidental dislodgement of guiding catheters.

## DISCUSSION

The coronary angiography and intervention trainer presented here could be valuable for novice and advanced interventional cardiologists. As virtual reality-based angiography simulation training has shown positive results, a physical system like the one we presented is likely to improve skills surrounding coronary angiography angulations and interpretation[2]. Technical skills that other non-digital PCI-focused trainers focused on can also be trained on our model[10,11]. Current non-digital trainers tend to be more reductive and focus on single tasks such as balloon exchange or two-dimensional wiring and interventions[10,11]. Currently available models do not meet the need to train complex situations such as complications and other high acuity low occurrence (HALO) procedures. Accordingly, interventional cardiologists may feel inappropriately prepared to deal with these events.

Simsiek et al. assessed the educational experience of interventional cardiologists in the US and Canada and found that even after dedicated training, less than half of the fellows felt comfortable performing various atherectomy techniques, implanting covered stents, fat embolization, or using embolic protection devices[12].

Interventions on porcine hearts for research, training, and educational purposes, as demonstrated in the Visible Heart Apparatus, are established as a valuable tool[13–16]Unfortunately, these methods are not easily accessible to trainees.

We provided instructions to make these procedures more accessible. Compared to a computer-based PCI simulator, which comes at a cost of ∼$55.000, we were able to compile a system for less than 50$. In contrast to the digitally simulated computer-based trainers, the model presented here allows any device to be used or tested. Properly cleaned used wires, balloons, catheters, and microcatheters can be trained and used for training to gain experience with these devices, as well as HALO procedures.

Furthermore, this model enables medical device prototype testing and the development or modification of interventional techniques. Eventually, data sets from CT scans of patients with difficult coronary lesions could be used to create 3D-printed lesions to incorporate into this model to have trainees perform a previous case completed by a different operator or to train robotic PCI devices.

## LIMITATIONS

While the simulator described here is reasonably realistic, some aspects lead to reduced fidelity of the system. As the guiding catheters are already in place, this system is not valuable for teaching catheter intubation or the difficulties in achieving and maintaining a good guide catheter position. There are no ECG or pressure curves on display in this setup. Learning to recognize ischemia on ECG or dampened and ventricularized pressure waveforms are important aspects of coronary intervention[17]. As the system is not pulsatile, there is no heart movement. Therefore, especially RCA interventions, which can be more difficult because of equipment movement, are not as realistic[18].

Porcine vessels are more compliant than diseased human coronary arteries and without coronary artery disease. Therefore, the haptic feedback of balloon inflation and wire manipulation will not be the same. Even though lesion characteristics can feasibly be modeled using 3-D printing, there is likely to be a difference. Similarly, the propensity to dissect or perforate differs between juvenile porcine arteries and diseased human coronary arteries.

Acquiring an appropriate heart specimen may pose difficulties. Ideally, large animal hearts from research facilities are used. Most commercially available hearts for human consumption are dissected and cannot be used. Hearts used for school dissection projects can be formalin-fixed and may show more tissue rigidity than is ideal for this setting.

## CONCLUSION

We report on how to build and use a low-cost simulator for coronary angiography and percutaneous coronary artery interventions. This model’s reusability and versatility make it a valuable tool in training interventional cardiologists. It could feasibly be used in any catheterization laboratory worldwide without requiring specific equipment for assembly and set-up. Dedicated training units with more specific preparation instructions and scenarios incorporating digital display simulation of arterial pressures and ECG are promising next steps to increase the simulator’s fidelity.

## Supporting information

Supplemental Digital Content 1

Supplemental Digital Content 2

Supplemental Digital Content 3

Supplemental Video Content 1

Supplemental Video Content 2

Supplemental Video Content 3

Supplemental Video Content 4

## ACKNOWLEDGMENTS

We thank the Visible Heart Laboratory staff and volunteers for their continuous engagement, which makes this such an inspiring place to work. We also thank Erik Donley for his work on the support structure.

## Data availability statement

Not applicable

## Ethics approval statement

Not applicable

## Patient consent statement

Not applicable

## Permission to reproduce material from other sources

Not applicable

## Clinical trial registration

Not applicable

