## Supplementary figures and images for "Enhancing Interventional Cardiology Training: A Porcine Heart-Based Coronary Intervention Simulator"

### Supplemental Digital Content 1

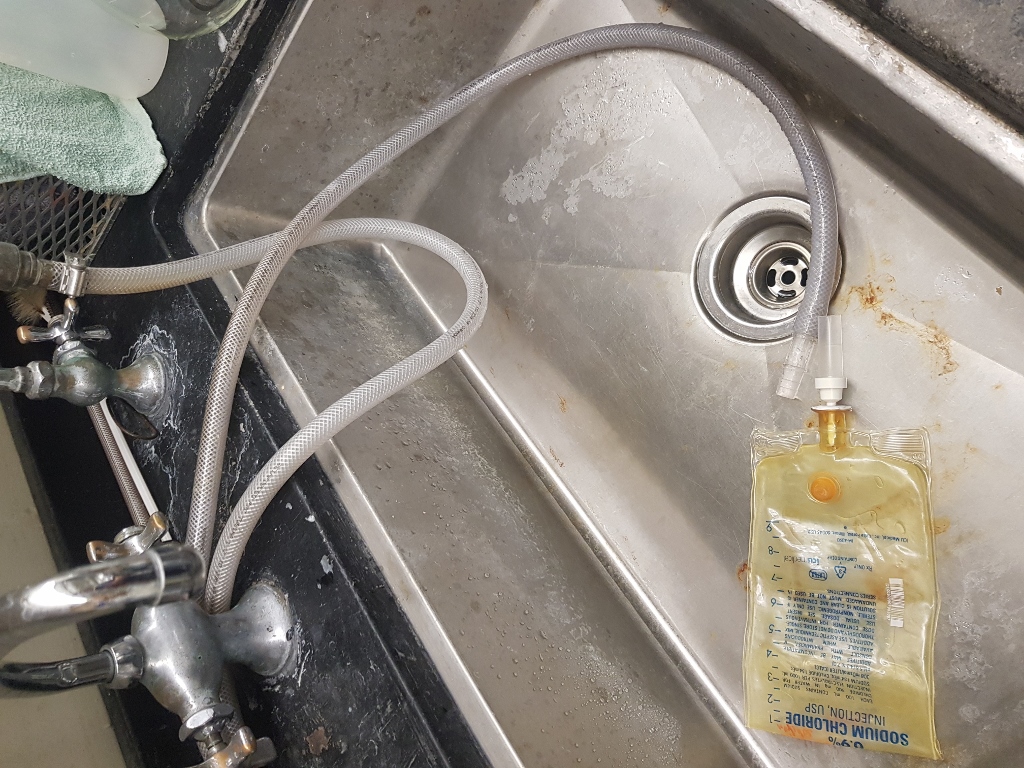

### Supplemental Digital Content 2

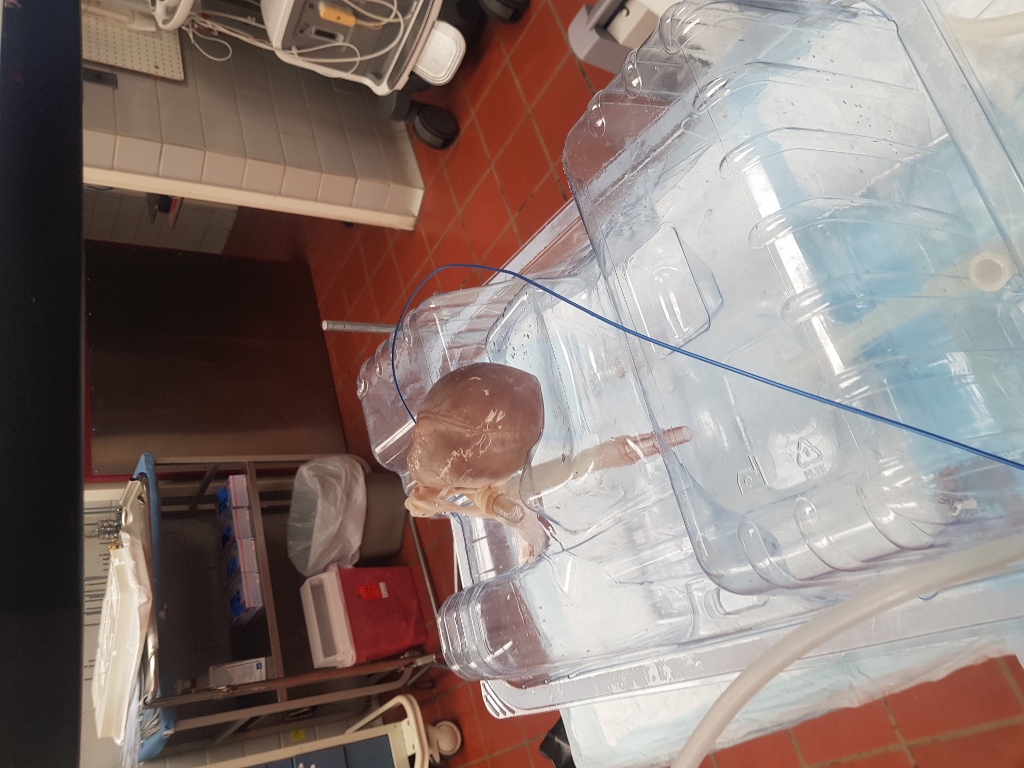

### Supplemental Digital Content 3

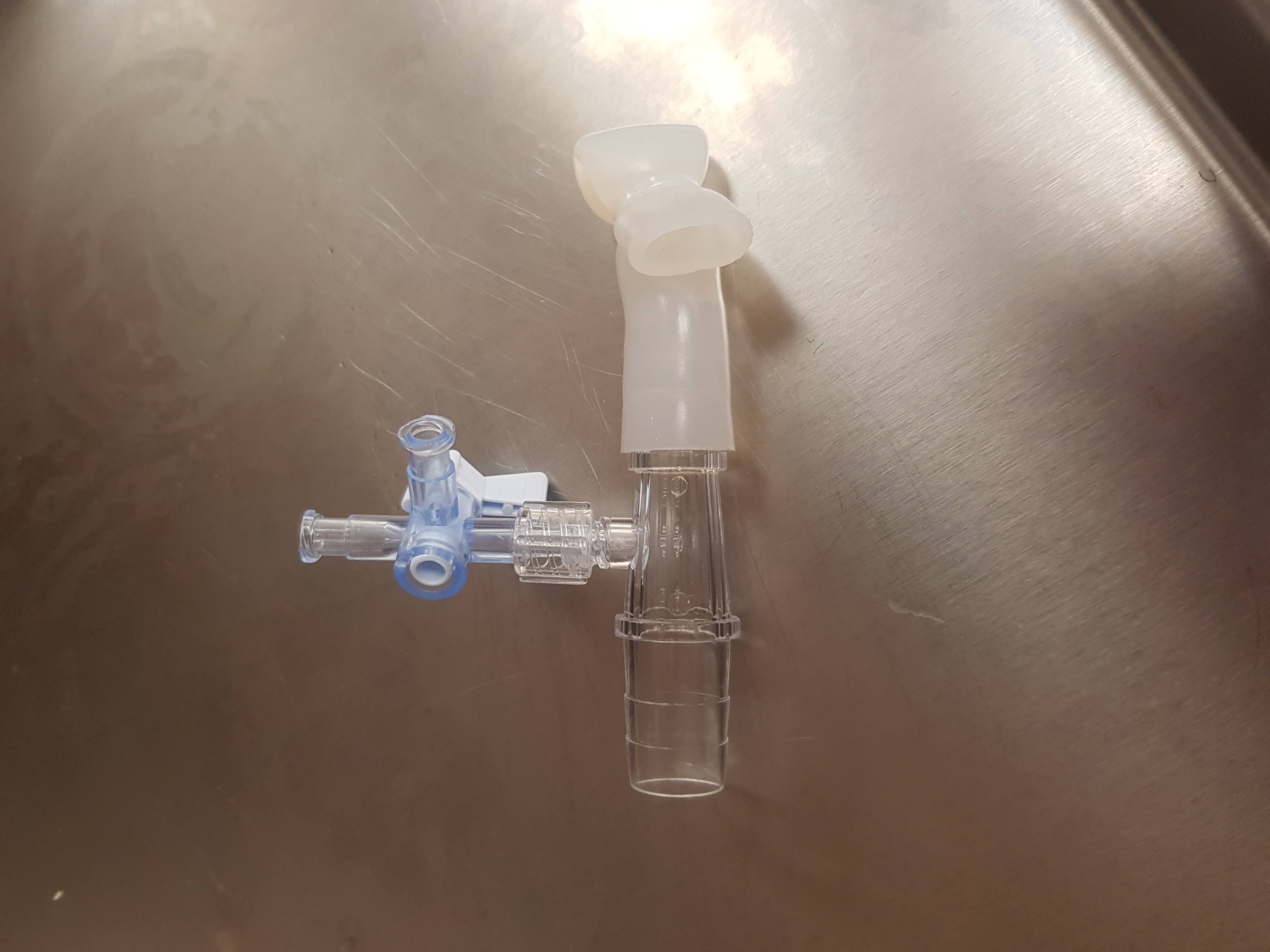
